# Shared HIV envelope-specific B cell clonotypes induced by a pox-protein vaccine regimen

**DOI:** 10.1101/2021.08.23.21262508

**Authors:** Kristen W. Cohen, Lamar Ballweber-Fleming, Michael Duff, Rachael E. Whaley, Aaron Seese, Daniel Imhoff, Zoe Moodie, Ollivier Hyrien, M. Juliana McElrath

**Affiliations:** Vaccine and Infectious Disease Division, Fred Hutchinson Cancer Research Center, Seattle, WA, U.S.A; Department of Medicine, University of Washington, Seattle, WA, U.S.A; Bristol Myers Squibb, Seattle, WA, U.S.A

**Author notes:** Address correspondence to: Ollivier Hyrien and M. Juliana McElrath, 1100 Fairview Ave. N., Seattle, WA 98109. and.

## Abstract

An effective HIV-1 vaccine will likely induce potent, broad neutralizing antibodies. No candidate vaccines have elicited these responses presumably because they fail to activate human B cell precursors that can affinity mature to generate broad neutralizing antibodies. To identify the B cell clonotypes that are elicited, we conducted in-depth analyses of the envelope-specific B cell repertoire in recipients of ALVAC-HIV vector (vCP2438) and bivalent subtype C gp120 protein (HVTN100). We observed high frequencies of envelope-specific IgG^+^ memory B cells with restricted immunogenetic diversity, relative to non-vaccine induced memory B cells, with preferential expansions of distinct variable genes but limited accumulation of mutations. Many envelope-specific clonotypes were shared across vaccinees, but did not overlap with the envelope-negative memory repertoire, within and across subjects. Single-cell sequencing of envelope-specific IgG^+^ memory B cells often revealed *VH1-2*02* and *VK3-20* sequence co-expression and in one case, contained a 5 amino acid CDRL3, the canonical signature of VRC01-class antibodies, confirming that these B cells are extremely rare but detectable. Our study provides evidence that immunogens play a critical role in selecting and restricting the responding B cell repertoire and supports the rational design of HIV vaccines targeting specific B cell lineages for induction of broadly-reactive neutralizing antibodies.

## Introduction

The HIV-1 epidemic is now in its fourth decade, and developing a highly effective preventive vaccine remains critical to end this global health crisis. The induction of broadly neutralizing antibodies (bnAbs) is widely thought to be a necessary component of a successful vaccine to prevent HIV-1 infection. This assumption is based on numerous preclinical studies indicating that passive administration of bnAbs can prevent SIV or SHIV infection following challenge (Balazs et al., 2014; Shingai et al., 2014), and it is further substantiated in humans, based on more recent findings that repeated IV infusions of the broadly neutralizing VRC01 mAb was associated with protection in humans against circulating VRC01-sensitive HIV-1 strains (Corey et al., 2021). However, no clinical vaccine strategies have thus far succeeded in reliably eliciting tier 2, including autologous strain-specific, neutralizing antibody responses. By contrast, approximately 30% of people living with HIV generate bnAbs within 2 years of infection, indicating that viral diversity pushes antibody maturation toward broad neutralizing activities (Doria-Rose et al., 2010; Hraber et al., 2014; Mikell et al., 2011; Piantadosi et al., 2009; Sather et al., 2012; Simek et al., 2009; van Gils et al., 2009). Thus in some cases, human B cells have the capacity to generate anti-HIV-1 bnAbs, and understanding the B cell events and molecular profiles required to induce bnAbs following immunization is a major priority in HIV development (Kwong et al., 2013).

When B cells differentiate from naïve to memory, they also undergo rounds of mutation and selection for higher affinity B cell receptors (BCR) and produce antibodies with increasing affinity to HIV-1 envelope (Env). B cells predicted to have the potential of evolving into mature B cells able to secrete bnAbs are found at extremely low frequencies in the naïve repertoire (Havenar-Daughton et al., 2018; Jardine et al., 2016a). Nevertheless, germline-targeting HIV vaccine designs have advanced into clinical trials to first prime germline bnAb precursors that can then be directed to affinity mature with boosts of sequentially more native-like HIV-1 envelopes (Briney et al., 2016; Jardine et al., 2013; Jardine et al., 2015; Jardine et al., 2016b; McGuire, 2019; McGuire et al., 2016; McGuire et al., 2013; Sok et al., 2016). Clinical trials of germline-targeting HIV immunogens aim to induce memory B cells expressing BCRs that share similarities to the unmutated common ancestors of known bnAbs. These trials rely on deep sequencing of the vaccine-induced BCR repertoire to evaluate the immunogenicity of the vaccine regimens. Few studies have investigated the molecular signatures of Env-specific BCR repertoires induced by HIV-1 vaccines and whether repeated immunizations with Env vaccines induce somatic hypermutation and features frequently associated with bnAbs (Basu et al., 2020; Easterhoff et al., 2017).

To address this issue, we examined the Env-specific memory B cell responses elicited in HVTN 100 (NCT02404311), a phase 1/2 randomized, double-blind, placebo-controlled clinical trial that evaluated the safety and immunogenicity of a clade B/C ALVAC-HIV (vCP2438) (months 0, 1) with bivalent subtype C gp120/MF59 (months 0, 1, 3, 6, 12) in HIV-seronegative low-risk populations South Africans (Bekker et al., 2018). Notably, the subtype C RV144-like vaccine regimen was subsequently tested in HVTN 702, a phase 2b-3 efficacy trial to evaluate HIV-1 prevention in populations at-risk in sub-Saharan Africa (Gray et al., 2021). The vaccine regimen did not induce nAbs (Bekker et al., 2018), and failed to protect against HIV-1 acquisition (Gray et al., 2021).

Here, we interrogated the BCR repertoire, the diversity of the Env-specific memory B cell repertoire, the extent of somatic hypermutation after repeated vaccination, the between-subject similarities and overlap between clonotypes and similarities to known bnAbs. Our findings of Env-specific BCR repertoires produced by conventional and ineffective Env gp120 immunogens in non-neutralizing HIV-1 prime-boost vaccine regimens offers a benchmark against which novel HIV-1 immunogens may be compared and provides additional context for the evaluation of germline-targeting immunogens.

## Results

### HVTN 100 evaluated a subtype C based modified RV144-like vaccine regimen

HVTN 100 was a phase 1 HIV-1 vaccine clinical trial of ALVAC-HIV vCP2438 including subtypes B and C Env immunogens plus MF59-adjuvanted bivalent subtype C 1086 and TV1 gp120 proteins. The study enrolled 250 participants in South Africa. (Bekker et al., 2018). Similar to the RV144 vaccine, the HVTN 100 regimen induced non-neutralizing Env-specific binding antibody responses that had the ability to mediate antibody-dependent cellular cytotoxicity (ADCC) along with polyfunctional Env-specific T cell responses (Bekker et al., 2018; Laher et al., 2020).

We performed in-depth analysis of the vaccine-induced peripheral Env-specific memory B cells in a subset of 14 out of 185 per protocol vaccine recipients enrolled in the HVTN 100 trial (Figure S1A). The subset was randomly selected among those with a positive serum binding antibody response to the vaccine-matched HIV envelope, 1086 gp120, in the bivalent protein immunogen. The demographics and vaccine-matched Env-specific binding antibody, antibody dependent cellular cytotoxicity (ADCC) and CD4+ T-cell responses of the subset were representative of those from the entire trial cohort (Figure S1B-S1E). No participants in the subset became HIV-infected during the study.

### Durability of Env-specific memory B cells 6 months after vaccination correlates with their expansion after boost immunization

To characterize the induction and maintenance of vaccine-elicited memory B cells, we evaluated, we evaluated the phenotype and magnitude of vaccine-matched subtype C 1086 gp120-specific memory B cells from vaccine recipients 6 months after the 4^th^ immunization, the day of 5^th^ immunization (month 12), and 2 weeks later (month 12.5) (Figure 1A and S1). Six months after the 4^th^ immunization, 1086 gp120-specific IgD^-^ memory B cells were low but detectable in all subjects (median 0.15%) and then increased significantly in response to the boost immunization (median 0.6%, Figure 1B p=0.0001). A minority of the total Env-specific IgD^-^ B cells were the IgM isotype (median 5.6%), which decreased to 1% after the 5^th^ vaccination (Figure 1C, p<0.0001). The majority of responding Env-specific IgD^-^ memory B cells were IgG^+^ at month 12 (median 64%). Two weeks after the 5^th^ vaccination (month 12.5), the proportion of Env-specific memory B cells that were IgG isotype increased significantly to a median of 79% (p=0.0047).

**Figure 1.**
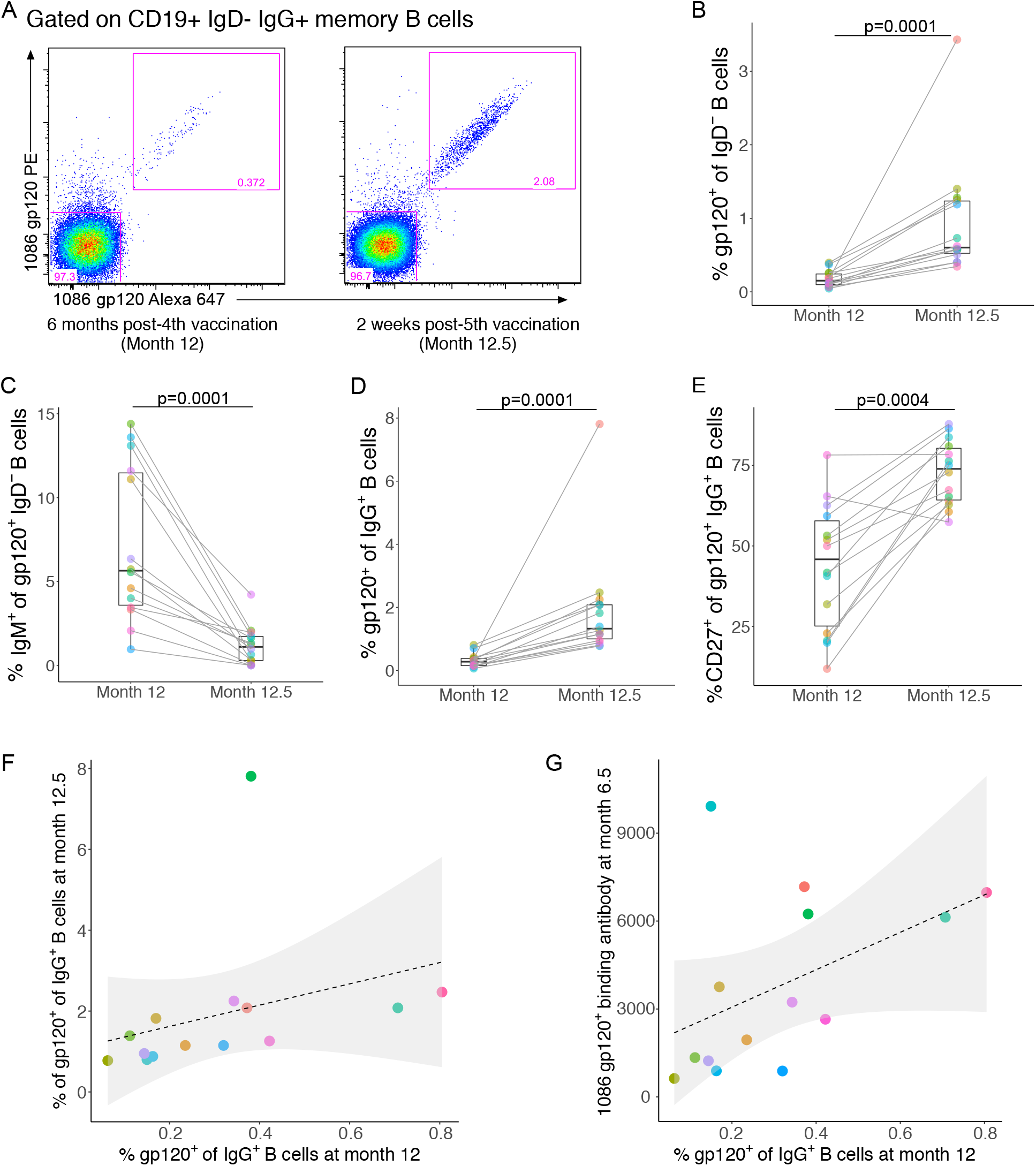
Env-specific memory B cells are detected 6 months after vaccination and expand significantly after boost immunization (n=14). Example of peripheral blood Env-specific memory B cells from one HVTN 100 participant gated on CD3^-^, CD14^-^, CD56^-^, CD19^+^, IgD^-^, and IgG^+^ live lymphocytes at A) month 12 (6 months post-4^th^ vaccination and day of boost) and month 12.5 (2 weeks after 5^th^ vaccination). Env-specificity is determined by double-staining with fluorescently-labeled 1086 gp120 probes. B) The frequencies of 1086 gp120^+^ IgD^-^ B cells at month 12 and 12.5. C) The proportion of gp120^+^ IgD^-^ B cells that were IgM^+^. D) The frequencies of 1086 gp120^+^ IgG^+^ B cells at month 12 and 12.5. E) The proportion of gp120^+^ IgG^+^ B cells that were CD27^+^. Boxes indicate the median (at line) and interquartile range. Statistical significance was determined using Wilcoxon signed rank test. P-values less than 0.05 were considered significant. The frequencies of 1086 gp120^+^ IgG^+^ B cells measured in PBMCs at month 12 correlated with the F) with the frequencies of 1086 gp120^+^ IgG^+^ B cells at visit month 12.5 and G) corresponding vaccine-matched serum binding antibody responses (net MFI by binding antibody multiplex assay) at month 6.5 (2 weeks post-4^th^).

The median frequency of gp120-specific IgG^+^ memory B cells was 0.28% at month 12 and increased 5-fold to 1.33% of IgG^+^ B cells at month 12.5 (Figure 1D, p=0.0001). Following the 5^th^ vaccination, the Env-specific IgG^+^ B cells also increased in CD27 expression (Figure 1E, p=0.0019) suggesting that the resting vaccine-specific memory B cells had lower CD27 expression 6 months post-vaccination, whereas the responding vaccine-specific B cells upregulated expression of CD27 and class-switched to IgG quickly after the boost.

Interestingly, the frequencies of gp120^+^ IgG^+^ B cells six months post 4^th^ vaccination/pre-boost (month 12) correlated significantly with the magnitude of gp120^+^ IgG^+^ B cells post-5^th^ vaccination (month 12.5) (Figure 1F; r=0.87, p=0.0027) and the magnitude of vaccine-matched serum binding antibody responses at peak (2 weeks post-4^th^ vaccination; Figure 1G; r=0.77, p=0.038). Thus, the frequencies of circulating Env-specific memory B cells were positively associated with the vaccine-elicited titers of Env-specific serum antibodies and predicted the magnitude of circulating B cells after boost.

### The vaccine-induced Env-specific BCR repertoire is narrower than the polyclonal non-vaccine selected memory repertoire

We explored the molecular immunogenetics of the vaccine-elicited memory B cell repertoire in order to identify characteristics shared by vaccine recipients. We isolated vaccine-specific B cells using fluorescent probes of the vaccine-matched 1086 gp120, which was the vaccine-matched Env with the highest serum antibody response. To characterize the B cell receptor (BCR) repertoire of Env-specific memory B cells, we sorted 1086 gp120^+^ and gp120^-^ (negative) IgG^+^ B cells two weeks post-5^th^ vaccination (month 12.5), which was the timepoint with the highest frequency of Env-specific memory B cells, and directly sequenced their *V_H_, V_K_* and *V_L_* immunoglobulin genes. We restricted cell sorting to IgG^+^ B cells because most vaccine-specific memory B cells were of IgG isotype. The majority of the gp120^-^ IgG^+^ cells are expected to be representative of the pre-vaccination polyclonal repertoire expanded in response to other infections or immunizations but not in response to the HIV vaccination. Thus, we sequenced the BCR of sorted 1086 gp120^+^ and gp120^-^ IgG^+^ B cells in parallel to compare the two repertoires.

We hypothesized that five doses of the Env-expressing vaccine regimen over one year drove the vaccine-specific B cells to undergo multiple rounds of expansion and selection, resulting in a narrower repertoire compared to the polyclonal non-vaccine specific memory B cell repertoire. Thus, we sought to compare diversity between the BCR repertoires of the 1086 gp120^+^ and gp120^-^ memory B cells (Figure S3). Since the diversity of a BCR repertoire is partly determined by the number of germline genes utilized to encode its heavy and light chains, we first assessed the diversity of the V genes encoding the gp120^+^ and gp120^-^ IgG^+^ repertoires using three indices of alpha diversity. The richness index is defined as the number of V genes encoding a given repertoire, indicated that the V regions of the heavy, kappa, and lambda chains of the vaccine-induced repertoire were encoded by fewer V genes than the non-vaccine induced repertoire (p < 0.001 in all cases; LMM adjusting for cell count and read count; Figure 2A-2C). The gp120^+^ IgG^+^ B cell repertoire had lower richness in 13 out 14 participants for the heavy chain (median of 55 versus 70 V_H_ genes), in 13 out 14 participants for the kappa chain (median of 32 versus 43 V_K_ genes), and in 14 out 14 participants for the lambda chain (median of 18 versus 30 V_L_ genes). Since the V gene richness may be sensitive to cell and read counts, we considered Shannon’s and Simpson’s indices as alternative metrics to assess diversity, both of which take evenness into consideration. Shannon’s index also suggested that the heavy, kappa, and lambda chains of the vaccine-induced repertoire were encoded by fewer V genes than in the non-vaccine induced repertoire (p < 0.001 in all cases; LMM adjusting for cell count and read count; Figure S2A). The Simpson index also indicated reduced diversity in the gp120^+^ repertoire of the kappa and lambda chains (p < 0.001; LMM adjusting for read count), but it did not reach significance for the heavy chain (p = 0.73; LMM adjusting for cell count and read count; Figure S2B). Taken together, these results suggest that, compared to the gp120^-^ IgG^+^ BCR repertoire, the heavy and light chains of gp120^+^ IgG^+^ BCR repertoire are encoded by a smaller set of V genes, and that within these V genes, some are preferentially selected and expanded. The observation of a focused vaccine-induced repertoire may be attributed to the preferential expansion of B cells with specific genetic signatures.

**Figure 2.**
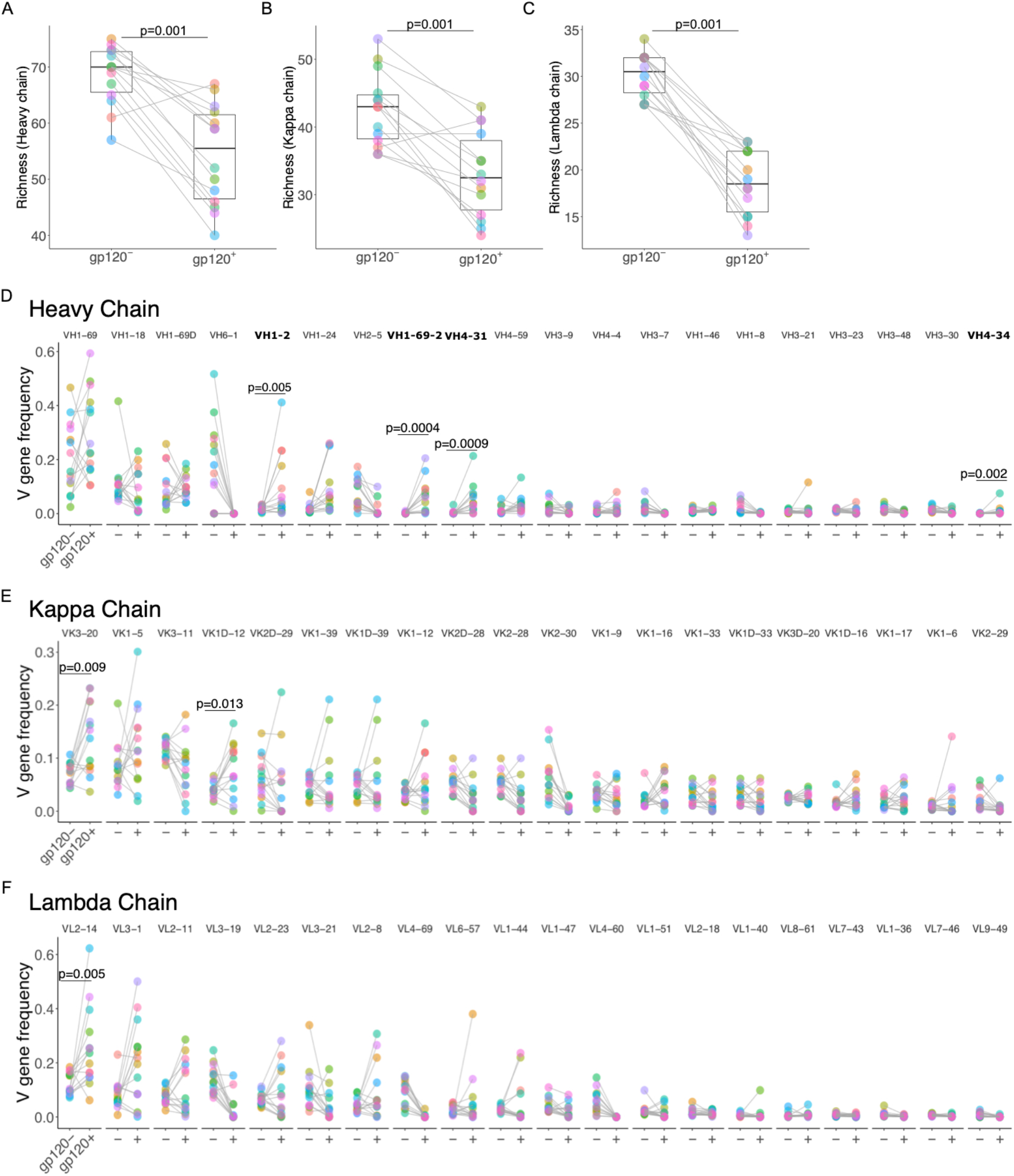
Diversity and V-gene distribution of paired vaccine gp120-specific and non-specific IgG^+^ B cell receptor repertoires (n=14). Diversity of V genes of gp120^+^ and gp120^-^ IgG^+^ B cell for *V_H_, V_K_*, and *V_L_* BCR sequences at month 12.5. A) heavy B) kappa and C) lambda chain diversity indices as calculated by Richness score among gp120-/+ IgG+ B cells. The paired frequencies per participant of 1086 gp120^+^ and gp120^-^ IgG BCR sequences expressing the top twenty: D) *V_H_*, E) *V_K_* or F) *V_L_* genes ranked by average frequency. D) Only the V_H_ genes that were significantly enriched in the gp120^+^ compared to gp120^-^ after adjusting for a FDR < 10% are indicated (in bold). All E) V_K_ and F) *V_L_* genes that were enriched in the gp120^+^ compared to gp120^-^ before adjusting for FDR are indicated; no V_K_ and *V_L_* genes were significant after controlling for FDR. Comparisons were conducted by Wilcoxon signed rank tests and show the unadjusted p values.

### In the gp120^+^ BCR repertoire, specific V genes were preferentially expanded across individuals

Ideally, a germline-targeting vaccine would induce an on-target B cell response consistent across vaccine recipients. To assess the similarity of the gp120^+^ BCR repertoire across vaccine-recipients, we measured the overlap between the sets of V genes that encoded each of the heavy, kappa, and lambda chains of gp120^+^ BCR from different individuals using the Jaccard index. The average Jaccard indices for the heavy, kappa and lambda chains of the gp120^+^ BCR repertoire were 0.64, 0.64 and 0.59, respectively. Thus, for all three chain types, on average about two thirds (66%) of the BCR in the gp120^+^ repertoire of two distinct vaccinees were encoded by the same pool of V genes, whereas the remaining one third (33%) of gp120^+^ BCR were encoded by distinct (i.e., non-overlapping) V genes. Although the value of two-third may under-estimate the overlap between vaccine-induced repertoires, it provides a benchmark for trials that target specific B cell lineages.

Since the Jaccard index indicated significant overlap in the use of V genes among the gp120^+^ B cells of distinct individuals, we next sought to determine which V genes were over-utilized by the gp120^+^ repertoire across participants. In evaluating the distribution of V gene families used by the gp120^+^ and gp120^-^ repertoires, we observed an immunodominance of the *V_H_1* family, in particular *V_H_1-69 and V_H_1-2*, and increased usage of the *V_H_4* gene family and decreased usage of the *V_H_6* and *V_H_2* gene families in the gp120^+^ compared to the gp120^-^ repertoires (Figure S3). In contrast, the distributions of kappa chain V gene families were similar between the repertoires and were dominated by *V_K_1*, *V_K_2* and *V_K_3* families (Figure S3). For lambda chains, the *V_L_4* family was enriched in the gp120^-^ repertoire and underrepresented in the gp120^+^ repertoire (Figure S3). We performed differential expression analysis to identify specific V genes unequally represented in the two repertoires (Figure 2D-2F; full list of comparisons can be found in the Supplement). For the *V_H_* BCR repertoire, we found that the *V_H_1-69-2, V_H_1-2, V_H_4-31 and V_H_4-34* genes were significantly expanded in the gp120^+^ repertoire compared to the gp120^-^ repertoire (unadjusted p values < 0.01 and FDR < 10%; Figure 2D). For the *V_K_* BCR repertoire, the *V_K_3-20* and *V_K_1D-12* genes were the most significantly enriched in the gp120^+^ relative to gp120^-^ BCR repertoire (unadjusted p=0.009 and 0.013, respectively; Figure 2E). For the *V_L_* BCR repertoire, the *V_L_2-14* gene was the most significantly enriched in the gp120^+^ compared to the gp120^-^ repertoire (unadjusted p=0.0052; Figure 2F). However, in contrast to the heavy chain, none of the light chain V genes were significant after controlling FDR for multiple comparisons. Additionally, multiple V genes were significantly under-represented in the gp120^+^ BCR repertoire and some V genes did not encode any of the gp120^+^ BCR isolated from the 14 vaccine-recipients (e.g., *V_H_6-1*).

### Evidence of limited somatic hypermutation in the gp120^+^ BCR repertoire

Since we observed selection and expansion of specific genes among the vaccine-specific B cells, we next asked whether the gp120^+^ IgG^+^ B cells had undergone substantial affinity maturation by month 12.5, after 5 vaccinations. When evaluating the frequency of nucleotide mutations of individual BCR sequences compared to their assigned germline, we found that the rates of mutation in the V gene segments of the heavy, kappa and lambda sequences were lower among the gp120^+^ compared to the gp120^-^ repertoires (Figure 3A-3C; p<0.001). For the *V_H_* BCR repertoire, the average percent mutation was 7.43% in the gp120^+^ repertoire and 8.4% in the gp120^-^ repertoire (Figure 3A). However, the rate of mutation observed in the gp120^+^ repertoire was very similar to the mutation rate observed after a 5^th^ vaccination of the RV144 regimen in the trial RV305 (avg 6.7%) (Easterhoff et al., 2017). The lower mutation rates among the vaccine-specific memory B cells after five vaccinations is indicative of limited affinity maturation, possibly due to limited or short-lived germinal center activity.

**Figure 3.**
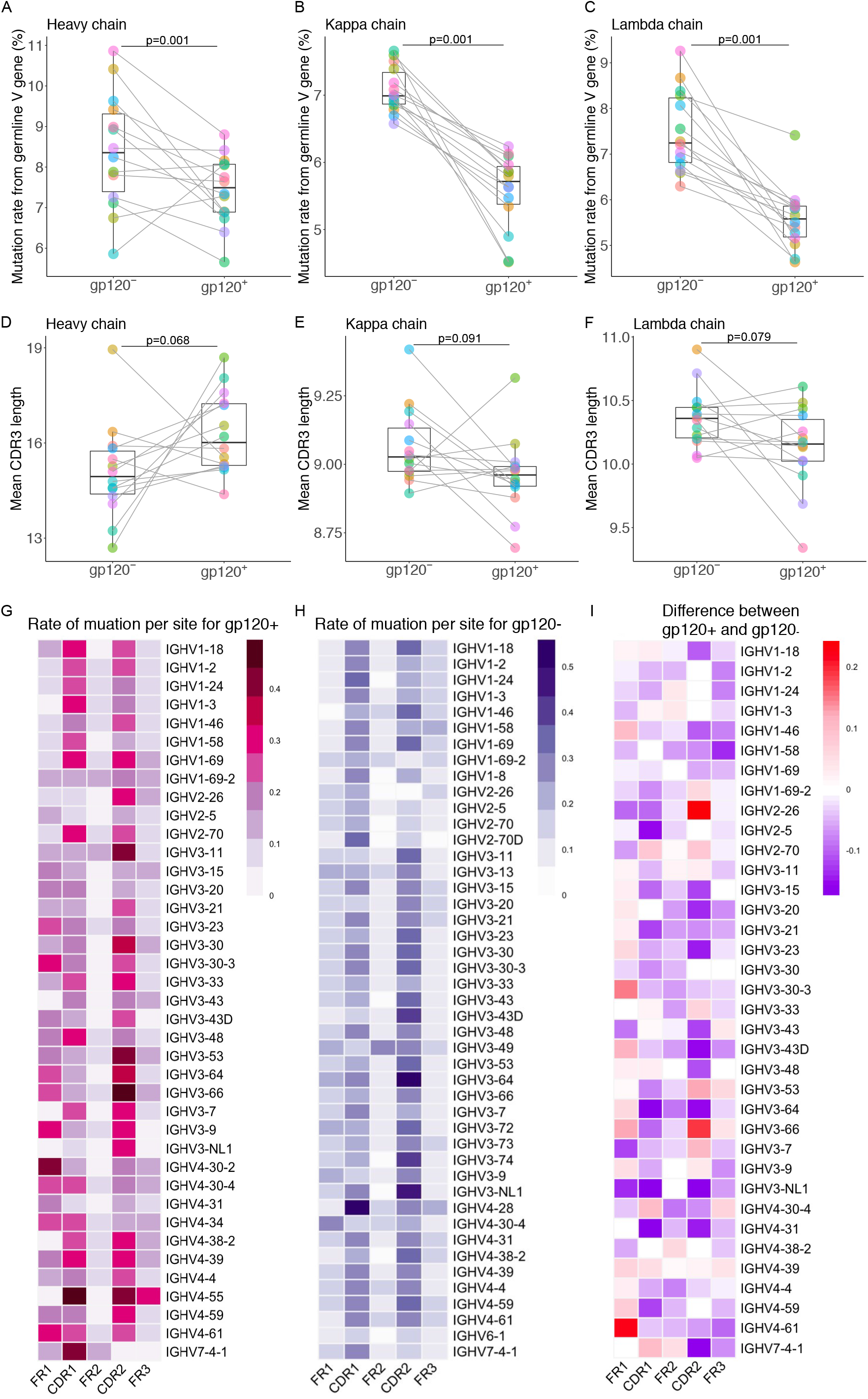
Mutation rate and CDR3 length among gp120^+^ and gp120^-^ *V_H_*, *V_K_* and *V_L_* BCR sequences at month 12.5 (n=14). Mean per subject percent mutation of the nucleic acid V gene sequences for the A) *V_H_*, B) *V_K_* and C) *V_L_.* Mean CDR3 lengths per subject for the D) heavy, E) kappa and F) lambda chains. Paired differences and corresponding P values were assessed using Wilcoxon signed rank tests. Mean amino acid (a.a.) mutations per site in the respective framework (FR) or complementary determining (CDR) regions of the *V_H_* sequence split out by V_H_ gene among G) gp120^+^ or H) gp120^-^ IgG^+^ B cells. I) the difference in a.a. mutation rate per V_H_ region between the gp120^+^ and gp120^-^ V_H_ sequences; cells in red indicate higher mutation rates in the gp120^+^ than in the gp120^-^ repertoires, and vice versa for those in blue.

### Vaccination did not induce antibodies with bnAb characteristics such as longer CDRH3 or shorter CDRL3 regions

Several anti-HIV-1 bnAbs have exceptionally long CDRH3 regions (e.g., PGDM1400, CDRH3 length=36; PGT121, CDRH3 length= 26). Their unusual lengths allow the bnAbs to reach into and between glycans on the HIV Env protein to make contacts with the amino acids (Sok et al., 2014). However, antibodies with long CDRH3 regions are uncommon in the naïve B cell repertoire and believed to be the result of rare VDJ recombination events. The CDR3 region is highly variable due to the random rearrangement of V(D)J gene segments and the addition and subtraction of nucleotides at the ends of these genes during somatic recombination. The average CDR3 lengths for the V_H_, V_K_ and V_L_ were not significantly different between the gp120^+^ and gp120^-^ repertoires (Figure 3D-3F). However, there was a trend toward longer CDRH3 lengths among the gp120^+^ compared to gp120^-^ BCRs (median=16.1 versus 14.9; p=0.068; Fig. 3D), but no significant differences in the frequency of BCRs with CDRH3 regions greater than 24 a.a. in gp120^+^ compared to gp120^-^ IgG^+^ B cells (p=0.30).

The average CDRL3 of both kappa and lambda chains is 9 a.a. long, but some HIV bnAbs, including VRC01-class bnAbs, are characterized by a 5 a.a. CDRL3 that is required to maintain a critical angle of approach to the CD4bs and prevent clashing with particular gp120 residues. These short CDRL3 are known to be rare in the naïve and memory B cell repertoires (Jardine et al., 2016a). We evaluated the CDRL3 length distribution of kappa and lambda BCR sequences from the gp120^+^ B cells compared to gp120^-^ B cells to assess expansion of light chains with short CDRL3 sequences. The median CDRL3 lengths among the kappa and lambda gp120^+^ BCR sequences, 9 and 10 a.a., respectively, were not significantly different from those of gp120^-^ B cells (Figure 3E and 3F). In addition, the frequency of light chains with a 5 a.a. CDRL3 sequence were rare and not different between the gp120+ and negative repertoires.

### Non-synonymous mutations concentrate in the complementary determining regions of V genes but don’t correlate with vaccine-specific V gene expansions

The rate of non-synonymous mutations in the gp120^+^ repertoire was explored by comparing the a.a. sequence of the V-gene encoded region of the light and heavy chains of each BCR sequence to that of their assigned germline gene. The average mutation rate per a.a. position in the FR1-3 and CDR1-2, by chain type and gene family Figure 3G-3I). When stratifying the analysis by framework (FR) and complementary determining regions (CDR; Figure 3G-3I), as expected, we observed higher accumulation of mutations in the CDRs compared to the FRs. The mutation rates in the *V_H_* genes continued to be lower among the gp120^+^ IgG^+^ B cells compared to gp120^-^ IgG^+^ B cells, even among the *V_H_* BCR sequences encoded by V genes that were specifically expanded in the gp120^+^ repertoire (i.e. *V_H_1-69-2, V_H_1-2, V_H_4-31 and V_H_4-34)*. Possible exceptions included the FR1 of BCR sequences encoded by the *V_H_4-61* family as well as the CDR2 of BCR sequences encoded by the *V_H_2-26* and *V_H_3-66* gene families which have accumulated more mutations in the gp120^+^ than in the gp120^-^ BCR repertoire (Figure 3I). However, these V_H_ genes were underrepresented in the gp120+ repertoire. Similarly, the rate of mutation in the *V_L_* genes was lower in gp120^+^ IgG^+^ B cells compared to gp120^-^ IgG^+^ B cells even when subdivided by V gene and region (Figure S4A and S4B). Overall, the CDR1 and CDR2 were the most mutated regions, followed by the FR1 and FR3 regions. The FR2 region was virtually unmutated. The amount of mutation varied by gene. Interestingly BCRs encoded by any of the *V_H_* genes found significantly over-expressed in the gp120+ repertoire were not among those with the highest mutation rates. In particular, the amount of mutations accumulated by the CDR1 and CDR2 regions of *V_H_1-2*02*-encoded BCR were relatively low compared to the CDR1 and CDR2 regions of BCR encoded by other gene families. Similar observations can be made for the kappa and lambda chains (Figure S4).

### Accumulation of mutations in V_H_1-2 gp120^+^ repertoire actually diverges the repertoire further away from representative V_H_1-2 bnAbs

Although vaccination induced BCRs with limited somatic hypermutations, we assessed whether it selected for BCR with mutations identical to those found in any *V_H_1-2*\*02-encoded bnAbs. Although encoded by a same V gene, the heavy chain of these bnAbs have accumulated distinct mutations, and we considered a representative subset of five V_H_1-2*02-containing bnAbs: VRC01 (Gristick et al., 2016), IOMA (Gristick et al., 2016), N6 (Huang et al., 2016), 3BNC117 (Scheid et al., 2011), and DH270.1 (Bonsignori et al., 2017). We calculated the Hamming distance between each *V_H_1-2*\*02-encoded read sequence (n=553 gp120^+^ BCR; n=35 gp120^-^ BCR) and the heavy chain of the five selected bnAbs, by region (FR1-3, and CDR1-2) and repertoire (gp120-vs. gp120+) (Figure 4). The results indicate that distances to the bnAbs were region-, bnAb-, and repertoire-specific. For instance, the distance to the CDR2 region of VRC01 was virtually identical between the gp120^-^ and gp120^+^ repertoires; however, the distances to the CDR2 region of the other four bnAbs were overall smaller in the gp120^-^ repertoire. Despite evidence for shared mutations with some bnAbs, the gp120^+^ BCRs encoded by the *V_H_1-2*\*02 gene remained overall distant to the selected ones. All FR3 read sequences were at least 9 a.a. mutations away from that of IOMA, 16 a.a. mutations from that of N6, 4 a.a. mutations from that of DH270.1, and 27 a.a. mutations from that of VRC-PG04. Overall, the mutation-selection profiles in each repertoire, by region and bnAb and suggests that mutations that accumulated in the gp120^+^ B cells moved the repertoire further away from the V_H_1-2*02 bnAbs (Figure 4).

**Figure 4.**
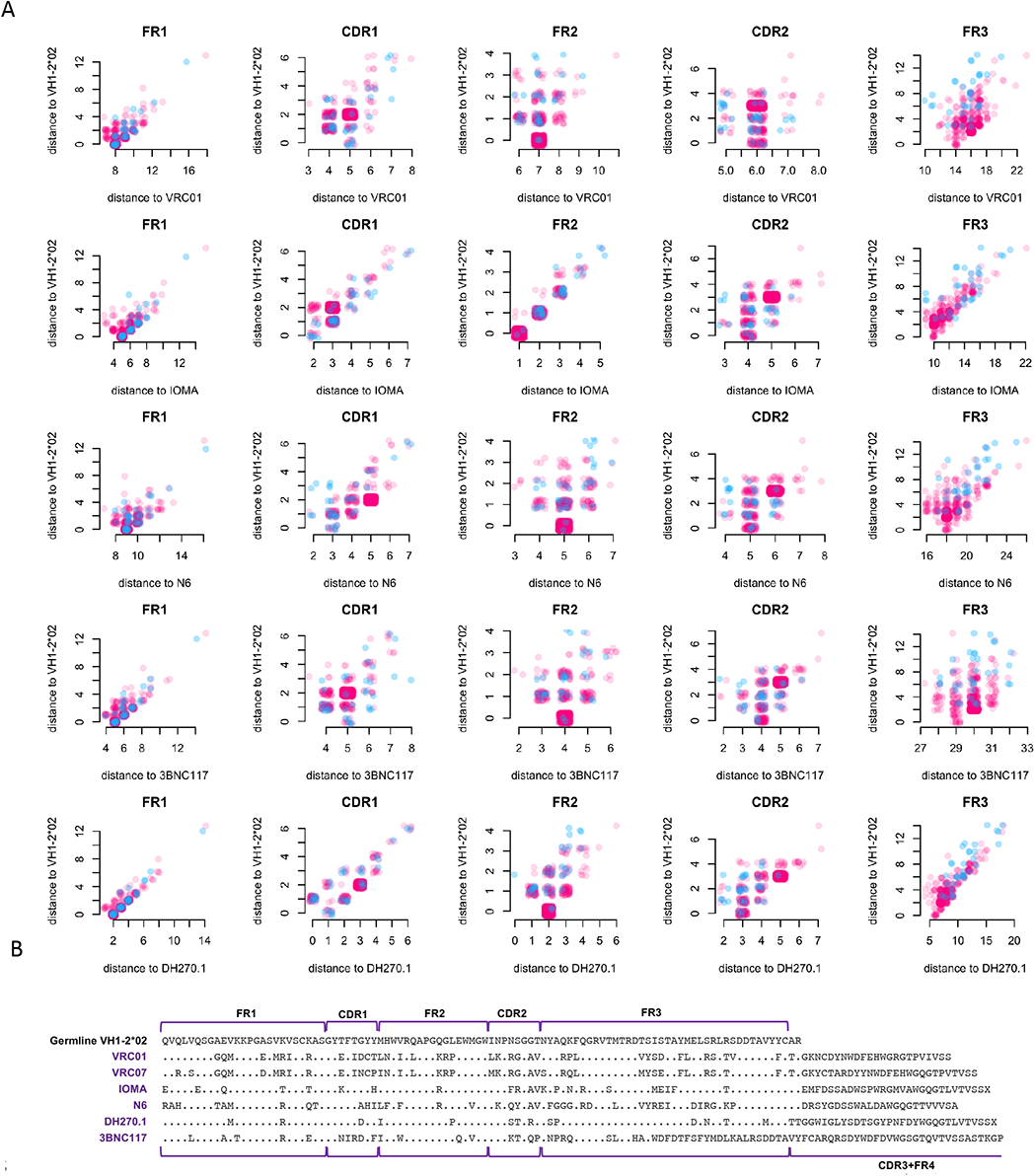
Mutations in the VH1-2*02-expressing Env-positive B cells do not lead to increased similarity to VH1-2*02 bnAbs. A) Among V_H_1-2*02-encoded gp120^+^ (pink) and gp120^-^ (blue) BCR sequences, the distance from the germline V_H_1-2*02 (y-axes) plotted against the distance from a subset of mature V_H_1-2-expressing bnAbs (from top to bottom rows): VRC01, IOMA, N6, 3BNC117, and DH270.1 per V gene framework (FR) or complementary determining (CDR) region. Each dot represents one BCR sequence; their color indicates the repertoire they were sampled from (red: gp120^+^; blue: gp120^-^). B) Comparison of the amino acid sequence of 6 bnAbs encoded by the VH1-2*02 germline V gene allele. Positions with a mutation relative to the germline VH1-2*02 sequence (top) are indicated.

### Shared vaccine-induced heavy chain clonotypes were detected across multiple subjects

We identified heavy chain clonotypes by clustering BCR sequences based on the similarity of the a.a. sequence of their CDRH3. The clustering process was blinded to the V and J gene calls of the sequences, their antigenic specificity (gp120^+^ vs. gp120-), and the participants from which they were sampled. Out of 13,381 unique sequences, we identified a total of 6,697 clonotypes. By inspecting the antigen-specificity of the BCR sequences assigned to identical clusters, we found that none of these clusters included sequences from both the gp120^+^ and gp120^-^ BCR repertoires (Figure 5A and 5D). In contrast, we observed that multiple clonotypes of unrelated origin were shared by several participants (Figure 5B-5D). Shared clonotypes are less likely to include BCR sequences from multiple participants if their clusters include few BCR sequences, and our analyses adjusted for cluster size (Figure 5C). Amongst the gp120^+^ clonotypes that consisted of at least 20 BCR sequences, at least 60% of them were shared by several participants (Figure 5C and 5D). In contrast, amongst the gp120^-^ clonotypes including at least 20 BCR sequences, we found that only about 15% of clonotypes were shared by at least two participants (Figure 5C and 5D). We conclude that the vaccine-induced repertoire was selected for in independent subjects indicative of convergent selection and evolution.

**Figure 5.**
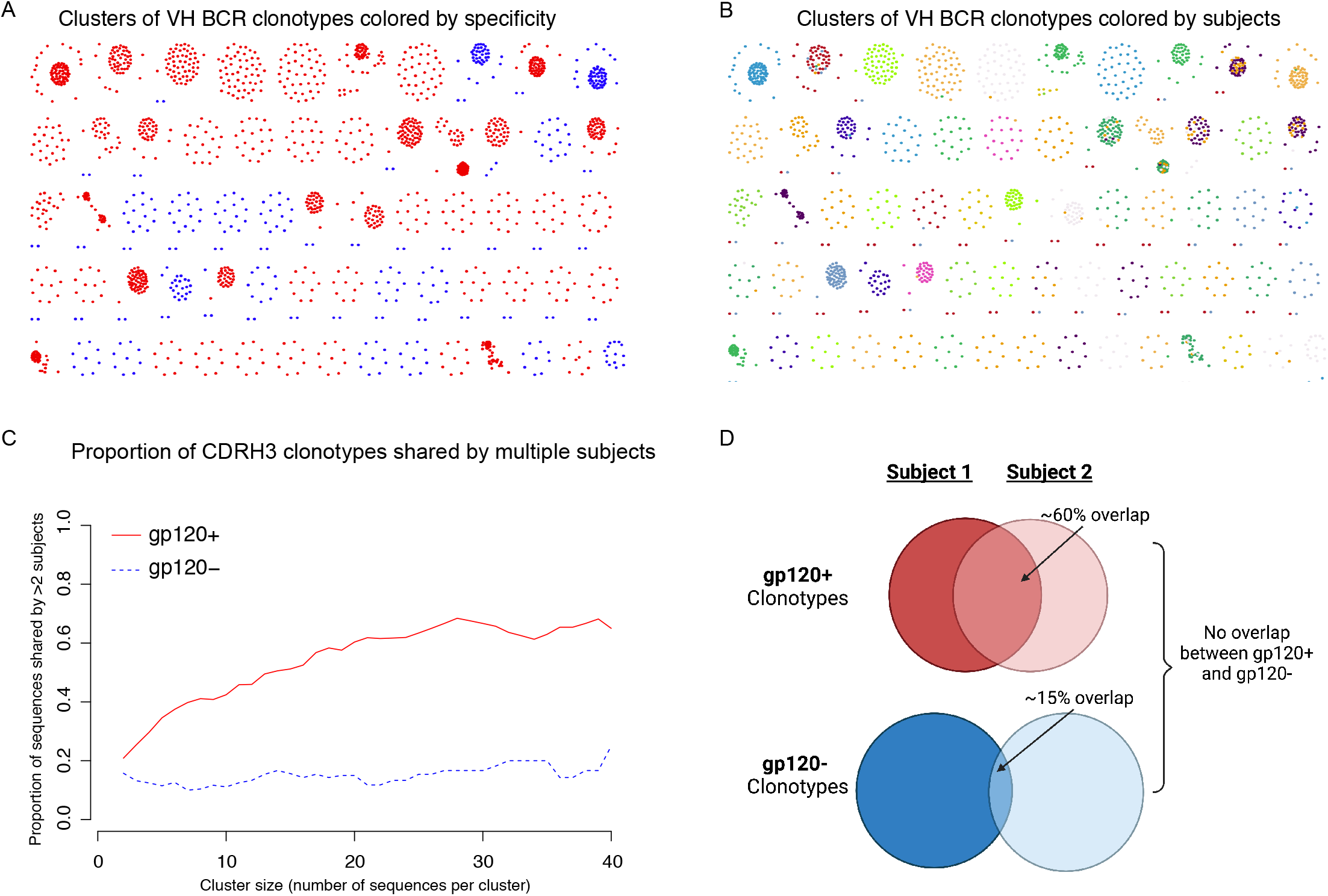
Shared B cell clonotypes were detected across subjects among the gp120+, but not gp120-BCR repertoires (n=14). Clonotypes were determined by clustering all CDRH3 sequences, regardless of class (gp120^+^ and gp120^-^) and subjects. A) Representative subset of the clusters of gp120^+^ and gp120-BCR V_H_ clonotypes, colored by the specificity of the BCR: gp120^+^ (red) and gp120^-^ (blue). B) Same representative subset of the clusters of gp120^+^ and gp120^-^ BCR V_H_ clonotypes, instead colored by subject that the sequence came from. C) The proportion of clonotypes shared by multiple subjects as a function of the size of the cluster of clonotypes for gp120^+^ and gp120^-^ B cells, respectively. D) A schematic of the average overlap in clonotypes between any two subjects for the gp120^+^ and gp120^-^ repertoires.

### High-throughput multiplexed next generation single-cell paired V_H_/V_L_ sequencing demonstrate detection of rare gp120^+^ IgG^+^ BCR clones

We sought to characterize the molecular characteristics of vaccine-induced BCRs with paired *V_H_* and *V_L_* BCR sequences, and selected the four vaccine-recipients with the largest expansions of *V_H_1-2*02* in their gp120-specific BCR repertoire analysis to evaluate the paired *V_H_* and *V_L_* BCRs. We performed single-cell sorting of 1086 gp120^+^ IgG^+^ memory B cells followed by next generation sequencing of their BCRs.

We obtained heavy and/or light chain BCR sequences from 1,912 gp120^+^ IgG^+^ B cells (1,364 heavy chain, 379 kappa chain and 568 lambda chain BCR sequences). A total of 339 gp120^+^ IgG^+^ B cells had paired heavy and light chain BCR sequences (159 kappa chains and 203 lambda chains, 23 cells registered both kappa and lambda chains). Even though the single-cell BCR sequencing analysis was limited to a subset of the participants analyzed in the bulk BCR sequencing analysis (n=4), it independently confirmed many of the patterns of gene usage observed in the bulk data set. In particular, we observed expansions of *V_H_1-2*02*, *V_K_3-20*, and *V_L_2-14* (Figure 6A and 6B). Furthermore, the analysis of B cells expressing paired heavy and light chains confirmed that the *V_H_1-2*02* heavy chain sequences were often paired with *V_K_3-20* light chains, which is noteworthy since the combination is a common motif among many VRC01-class BCRs. Specifically of the 1,364 cells with heavy chain sequences, 181 contained a *V_H_1-2*02* and of these, 38 of these were paired with a light chain. The average length of the amino acid sequence of the CDR3 of the light chain of the 38 sequences with a paired *V_H_1-2*02* heavy chain V gene was 8.6 for kappa and 10.3 for lambda. Two light chains paired with *V_H_1-2*02* had five amino acid-long CDRL3s (*V_K_3-20*, QHMYT and *V_K_1-5*, LPFRV), with one being a *V_K_3-20*. These data confirm that Env-specific B cells with molecular signatures associated with VRC01-class of bnAbs can be induced by a non-neutralizing gp120-based prime-boost vaccine but these B cells are extremely rare (∼ 2.6% of V_H_1-2*02 gp120^+^ B cells or 0.29% of gp120^+^ B cells) and their relevance is unknown at this point, except that it provides a benchmark to which VRC01 germline-targeting vaccines can be prepared.

**Figure 6.**
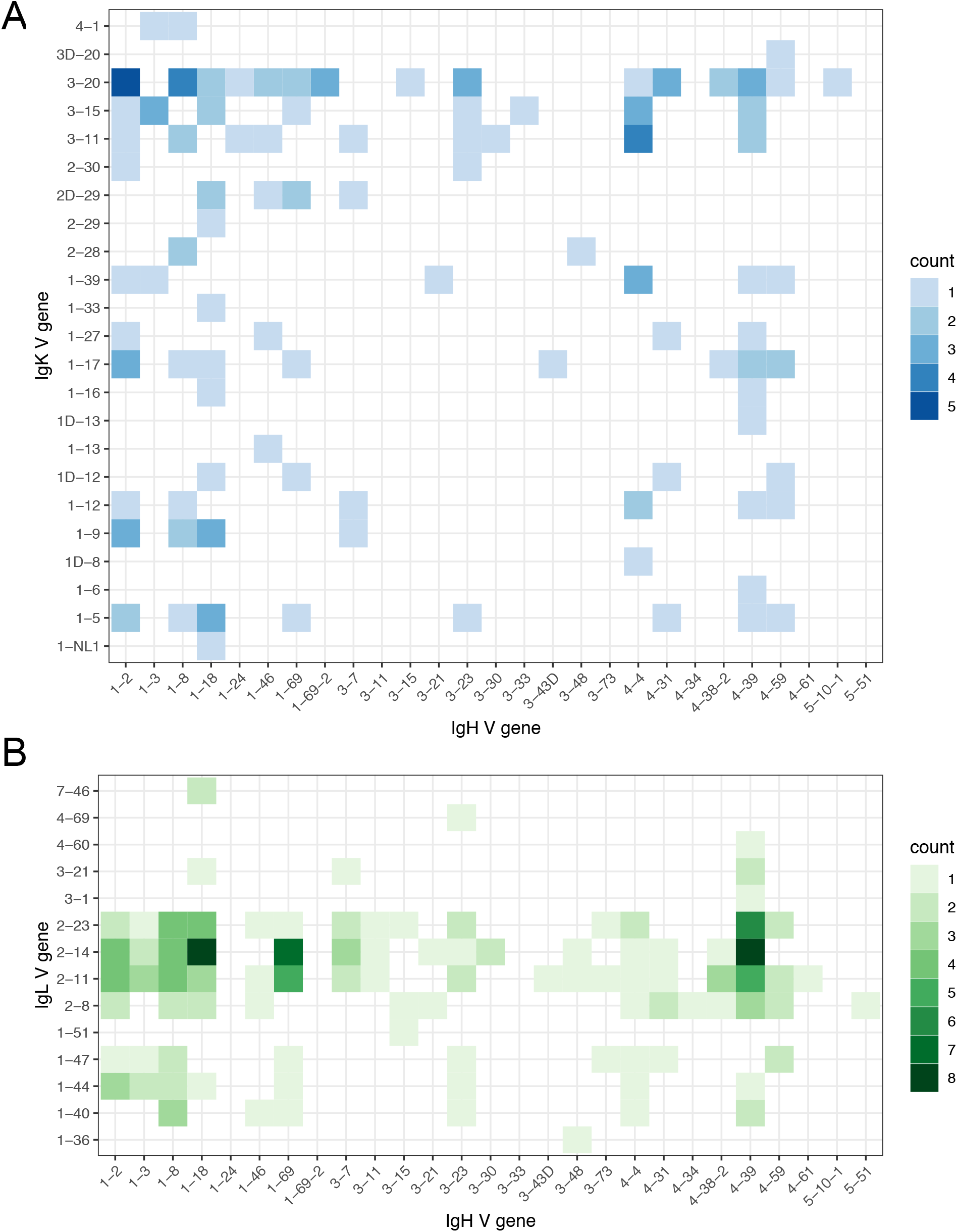
Single-cell BCR sequencing identifies immunodominant heavy and light chain pairs. Counts of paired heavy and light chain sequences from single-cell gp120^+^ IgG^+^ B cells from 4 vaccinated HVTN 100 subjects with expanded *V_H_*1-2*02 gp120^+^ B cells for A) *V_H_/V_K_* and B) *V_H_/V_L_*, respectively.

## Discussion

Most effective vaccines may protect against pathogens through multiple mechanisms, and antibodies may contribute through both neutralizing and non-neutralizing effector activities. Despite tremendous effort spanning many decades, an efficacious HIV vaccine continues to elude the field. However, recent technical advances, allowing for improved Env immunogen design and engineering, and tracking of the resultant humoral responses have provided renewed enthusiasm. Here, we have detailed the phenotypic and molecular characteristics of Env-specific B cells generated by a vaccine regimen in a pivotal trial (HVTN 100) that supported the decision to advance to a phase 2b/3 vaccine efficacy trial (HVTN 702). Although the vaccine regimen tested in HVTN 100 was not found to prevent HIV infection, we observed that the pox/protein prime-boost regimen induced high frequencies of class-switched Env-specific memory B cells that they were durable up to 6 month post 4^th^ vaccination. Of note, the original 4 vaccination RV144 vaccine regimen waned in efficacy after 6 months post 4^th^ vaccination (Robb et al., 2012). Consequently, a 5^th^ vaccination was added to the HVTN 100 regimen at 6 months post-4^th^ vaccination in order to boost and extend vaccine-induced immunity. Our findings indicate that the 5^th^ vaccination rapidly increased the Env-specific memory B cells frequency (∼5-fold), CD27 expression and IgG isotype, which together support the finding that the vaccine regimen, including the 5^th^ vaccination/boost, effectively induced and expanded HIV gp120-specific memory B cells. Moreover, the frequencies of gp120^+^ IgG^+^ B cells prior to the 5^th^ vaccination correlated with the frequency of gp120^+^ IgG^+^ B cells 2 weeks later and peak serum antibody responses, highlighting the direct relationship between memory B cells in circulation and bone marrow plasma cells, which secrete the serum antibodies.

In order to characterize the immunogenetics and clonotypes of vaccine-induced Env-specific memory B cells, we conducted next generation sequencing of the BCR repertoires of gp120-specific (gp120^+^) and non-vaccine specific (gp120^-^) IgG^+^ memory B cells from the 14 subjects 2 weeks after the 5^th^ vaccination. We found that the gp120^-^ specific memory BCR repertoire was largely of the V_H_1 gene family. This is similar to a recent study of early circulating plasmablast and long-lived (>8 months) bone marrow plasma cell repertoires from a RV144-like DNA/gp120 vaccine regimen (Basu et al., 2020). In particular, in the gp120^+^ repertoire, *V_H_1-69* and *V_H_1-2* were immunodominant, which have been previously observed among Env-specific repertoires induced both by HIV vaccination and infection (Basu et al., 2020; Huang et al., 2004; Smith et al., 2018). The gp120-specific repertoire was less diverse than the gp120^-^ memory repertoire, which is consistent with the hypothesis that the gp120^-^ repertoire comprised of non-vaccine induced B cells with diverse specificities in contrast to the gp120^+^ vaccine-induced repertoire. Interestingly, the estimates of BCR beta diversity further suggested that about two-thirds of the heavy and light V genes in the gp120^+^ repertoires were shared between any two subjects. In fact, multiple V_H_ genes were significantly expanded specifically in the gp120^+^ repertoire compared to the gp120^-^ memory repertoire consistently across subjects, including *V_H_1-69-2, V_H_1-2, V_H_4-31* and *V_H_4-34*. The V_H_1-2 expansion is particularly interesting as it is a defining feature of the VRC01-class of bnAbs and its germline sequence encodes for many of the residues that are critical for binding envelope and neutralizing activity (Kwong and Mascola, 2012; West et al., 2012). Indeed, novel immunogens have been engineered to engage and activate VRC01-class bnAb precursors and are now being tested in clinical trials for their ability to expand VRC01-like B cells (CD4bs-specific with VH1-2*02,03 or 04 paired with a light chain with a 5 a.a. CDRL3) (Jardine et al., 2013; Jardine et al., 2016a; McGuire et al., 2013). Thus, the consistent expansion of gp120^+^ VH1-2*02 memory B cells by vaccination with a gp120-based HIV vaccine suggests these B cells can be readily expanded by vaccination and that VRC01 germline-targeting vaccines will need to further focus the response to avoid off-target responses.

We also identified *V_H_4-34* as one of the V gene segments statistically significantly over-represented in the vaccine-induced repertoire. Previous studies have reported that *V_H_4-34* BCRs may be associated with autoimmune disorders and autoreactivity (Bhat et al., 2002) and enriched among highly mutated BCRs of HIV-1-infected individuals (Roskin et al., 2020). Although over-represented in the vaccine-induced repertoire, the frequency of *V_H_4-34* BCR remained low in our cohort (mean=0.39% vs 0.027% in the gp120^+^ vs gp120^-^ repertoire). In addition, we observed that the FR1 of *V_H_4-34* BCR tended to be more mutated than the FR1 of those encoded by other V gene segments. Somatic hypermutations also accumulated in their CDR1, but mutations were overall rare in the FR2, CDR2 and FR3. This mutational profile is consistent with previous reports that implicated FR1 in the antigen binding by *V_H_4-34* (Li et al., 1996). Taken together, these results suggest that vaccination might have partly recapitulated HIV exposure in inducing *V_H_4-34* clones; however, while the FR1 and CDR1 were the most prone to somatic hypermutation, we did not find evidence of higher mutation rates of codon 26 in the gp120^+^ than in the gp120^-^ repertoire, unlike previously observed in a cohort of HIV-1 infected patients (Roskin et al., 2020). The enrichment of certain V_H_ genes among antigen-specific B cells has also been observed in the humoral response to COVID-19 and SARS-CoV-2 vaccination (Chen et al., 2021b; Kim et al., 2021). It is not unexpected that the vaccine-induced repertoire would be focused and is likely attributable to the preferential expansion of B cells with specific V gene characteristics in response to vaccination.

Since we observed clear selection and expansions of specific gene families among the vaccine-specific B cells, we were surprised to discover that the gp120^+^ IgG^+^ B cells had undergone limited somatic hypermutation after 5 vaccinations. The rate of mutation in the gp120^+^ memory V_H_ repertoire was similar to previous reports of RV305 vaccine recipients who had received 5 doses of the RV144 vaccine regimen (Easterhoff et al., 2017). However, the mutation rate remained significantly lower than the gp120^-^ memory repertoire. Our data suggest that despite receiving five immunizations, vaccine-responding B cells display a reduced level of affinity maturation as compared to B cells from natural infection with other pathogens, as indicated by the gp120^-^ repertoire. The rate of non-synonymous mutations in the gp120^+^ repertoire was further explored by comparing the amino acid sequence of the V-gene encoded region of the light and heavy chains of each BCR sequence to that of their assigned germline gene. We found that the non-synonymous mutations accumulated in the complementary determining regions of V genes but did not correlate with the V genes that had preferentially expanded. This suggests that although some V genes were preferentially expanded, they did not undergo sustained germinal center-associated maturation. Although the vaccination-induced BCRs had limited somatic hypermutations, we assessed the tendency of *V_H_1-2*02*-encoded gp120^+^ BCRs to share mutations with those found in *V_H_1-2*02*-encoded bnAbs and found that the limited somatic hypermutation in the gp120^+^ B cells appeared to diverge further from the *V_H_1-2*02* bnAbs.

To further understand the similarity of B cell responses across subjects, we evaluated CDRH3 clonotypes, identifying 6,697 unique clonotypes, none of which overlapped between the gp120^+^ and gp120^-^ repertoires even within individuals. The lack of overlap between the gp120^+^ and gp120^-^ clonotypes provides further support that the gp120^-^ pool of memory B cells is representative of the non-vaccine induced repertoire. Among the vaccine-specific clusters (gp120^+^), we identified multiple publicly shared vaccine-induced clonotypes across multiple subjects. Public clonotypes have been described in response to HIV-1 infection, including Env-specific V_H_1-69 and V_H_1-2 clonotypes (Setliff et al., 2018; Smith et al., 2018), as well as to other pathogens such as SARS-CoV-2 vaccination and infection, including neutralizing and non-neutralizing clonotypes (Chen et al., 2021a). These observations further confirm the role of the immunogen, including recombinant gp120, in selecting the repertoire of the responding B cells with shared molecular characteristics across subjects.

Lastly, independent high-throughput multiplexed next generation single-cell paired V_H_/V_L_ sequencing confirmed the enrichment of vaccine-induced gp120^+^ B cells co-expressing the canonical VRC01-class V genes V_H_1-2*02 paired with V_K_ 3-20 light chain. Because inferred germline versions of VRC01-class bnAbs do not bind recombinant gp120 *in vitro*, it is hypothesized that current HIV-1 Env-based immunogens would not activate these BCRs. glVRC01-class targeting immunogens aim to expand B cells with BCRs containing V_H_1-2*02 (*03 or *04) alleles paired with a short (5 a.a.) CDRL3. However, only 1 of these V_H_ 1-2*02/V_K_ 3-20 expressing gp120^+^ B cells had a light chain with a 5 a.a. CDRL3, and the resulting monoclonal antibody, FH1, was found to not bind the CD4bs. Therefore, rare vaccine-induced gp120^+^ clones with VRC01-class molecular characteristics can be detected, but it is critical to know the epitope specificity to ascertain whether the without the corresponding epitope specificity their presence could be misinterpreted.

A major hurdle in driving development of HIV-specific bnAbs by vaccination is the ability to activate specific B cell lineages and induce affinity maturation through multiple rounds of mutation and selection. We have shown that the pox-protein vaccine strategy induced limited somatic hypermutation but did engage a restricted repertoire of vaccine-reactive B cells. Understanding the lineages of B cells recruited and expanded by non-neutralizing gp120-based HIV vaccine strategies will help inform interpretation of upcoming germline-targeting HIV vaccine trials and guide vaccine strategies.

## Materials and Methods

### Study design, population, products, and procedures

HVTN 100 was a randomized, controlled, double-blind study phase 1-2 clinical trial, which enrolled 252 healthy HIV-uninfected 18- to 40-year-old participants at 6 sites in South Africa. The trial was registered with the South African National Clinical Trials Registry (DOH-27-0215-4796) and ClinicalTrials.gov (<u>NCT02404311</u>). The investigational products were ALVAC-HIV (vCP2438) and MF59-adjuvanted bivalent subtype C gp120. ALVAC-HIV (vCP2438) (viral titer nominal dose of 10^7^ 50% cell culture infectious dose) expressed the envelope gp120 of the subtype C ZM96.C strain with the gp41 transmembrane sequence of the subtype B LAI strain, as well as *gag* and *protease* from the subtype B LAI strain. Bivalent subtype C gp120 was a combination of 100 μg each mixed with the MF59 adjuvant, a squalene oil-in-water emulsion. Placebo for ALVAC-HIV was a mixture of virus stabilizer and freeze-drying medium reconstituted with 0.4% NaCl, and placebo for the bivalent subtype C gp120 was 0.9% NaCl for injection. Participants were randomized in a 5:1 vaccine:placebo ratio to receive ALVAC-HIV at months 0 and 1 followed by 3 administrations of ALVAC-HIV along with bivalent subtype C gp120/MF59 at months 3, 6, and 12, or to receive placebo throughout, all by intramuscular injection. The research ethics committees of the University of the Witwatersrand, the University of Cape Town, the University of KwaZulu-Natal, and the Medical Research Council approved the study. All participants gave written informed consent in English or their local language (Setswana, Sotho, Xhosa, or Zulu). Additional detail about the HVTN 100 study design, eligibility criteria, participants and their baseline characteristics, randomization, blinding, study products is available in previous reports (Bekker et al., 2018).

### Binding antibody multiplex assay

HIV-1-specific IgG binding antibody responses were measured with 1:200 dilutions of serum for presence of IgG that was able to bind to the gp120 immunogens by an HIV-1 binding antibody multiplex assay as described previously (Tomaras et al., 2008; Yates et al., 2018).

### Intracellular cytokine staining assay

CD4+ T-cell responses to HIV vaccine insert-matched peptides were measured by intracellular cytokine staining. The assay detects the production and accumulation of cytokines on inhibition of intracellular transport after brief cell stimulation. Cryopreserved peripheral blood mononuclear cells were thawed, rested overnight, and stimulated with a peptide pools representing the 108.c gp120 ALVAC insert ZM96 gp120 and dimethyl sulfoxide (DMSO; negative control) or *staphylococcal enterotoxin B* (SEB; positive control) in the presence of costimulatory antibodies (CD28 and CD49d), and intracellular transport inhibitors brefeldin A and monensin for 6 h at 37°C. Next, cells were washed and incubated with EDTA (edetic acid) overnight at 4°C, then stained with a 16-colour panel (Moncunill et al., 2015), acquired on a BD LSRII flow cytometer (BD Biosciences, San Jose, CA, USA), and analyzed using FlowJo version 9.9.4 (BD, Franklin Lakes, NJ, USA).

### B cell phenotyping and sorting

Cryopreserved peripheral blood mononuclear cells (PBMCs) were thawed and transferred to warm thaw media supplemented with benzonase [RPMI, 10% fetal bovine serum (FBS), 1% L-Glutamine, 1% penicillin-streptomycin, benzonase 50 U/mL] followed by two additional washes with thaw media. Samples were rested overnight at 37°C 5% CO_2_ and centrifuged at 250 x g for 10 minutes before transferring to a 96-well U-bottom plate for staining. Cells were pelleted for 3 minutes at 750 x g and resuspended in Live/Dead marker (AViD, Invitrogen) diluted in 1x PBS. After a 15 to 20-minute incubation at 4°C, Live/Dead stain was washed off with PBS and PBMCs were incubated for 8-15 minutes at 4°C in anti-human CD4 (BD Biosciences) diluted in stain buffer (PBS, 10% FBS) to block non-B cell receptor binding of antigens to cells via the Envelope CD4-binding site. Two probes were prepared in advance at a molar ratio of 4:1, protein to streptavidin (SA) using HIV-1_1086_ Env gp120 conjugated to phycoerythrin (PE)-SA (BD Biosciences) and AlexaFluor647-SA (Invitrogen). The two tetramers were diluted in stain buffer and incubated with cells for 30 minutes at 4°C. Samples were washed with stain buffer, pelleted and resuspended with the following antibody panel diluted in stain buffer for 15-20 minutes at 4°C: anti-CD3 BV510 (BD Biosciences, UCHT1), anti-CD14 BV510 (BD Biosciences, MfP9), anti-CD56 BV510 (BD Biosciences, NCAM16.2), anti-CD19 PE-Cy7 (BD Biosciences, SJ25C1), anti-CD27 BV605 (BD Biosciences), anti-IgD FITC (BD Biosciences), anti-IgM PE-Dazzle594 (BioLegend, MHM-88), anti-IgG BV450 (BD Biosciences), and anti-CD38 PerCP-Cy5.5 (BD Biosciences, HIT2), anti-CD21 BV711 (BD Biosciences), anti-CD16 (BD Biosciences), anti-CD32 (BD Biosciences), anti-CD64 (BD Biosciences), and Mouse IgG. The samples were washed and resuspended in stain buffer for collection on a BD FACS Aria II (BD Biosciences). Two populations (HIV-1_1086_ Env gp120 positive and HIV-1_1086_ Env gp120 negative) were bulk sorted into separate FACS tubes containing lysis buffer [DNA suspension buffer (Teknova), 10% BSA, 2.5% RNAseOut] using the following gating strategy: singlets, lymphocytes, Live/Dead-, CD3-, CD56-, CD14-, CD19+ IgD-, IgG+, 1086 gp120+ or 1086 gp120-.

Four of the original subset were selected for further single-cell analysis and were thawed and stained as described above with the following modifications: IgD BV650 (BD Biosciences, IA6-2) and IgG BV786 (BD Biosciences, G18-145). Cells that were 1086 gp120+ were single cell sorted using the gating scheme described above into PCR plates containing lysis buffer. Plates with sorted cells were sealed and frozen on dry ice prior to transferring to −80°C for storage.

### Next generation BCR repertoire sequencing

Total RNA from HIV-1_1086_ Env gp120 positive and HIV-1_1086_ Env gp120 negative bulk-sorted cells were extracted using the RNeasy Micro Kit (Qiagen) according to the manufacturer’s protocol. Random primed cDNA synthesis was carried out with 10 μL of total RNA using SuperScript III Reverse Transcriptase Kit (Invitrogen/Thermo Fisher) per the manufacturer’s protocol.

For the heavy chain immunoglobulin transcripts were amplified using three separate primary previously published PCR primer pools (G1-G3, see Supplemental for list of primers) (Scheid et al., 2011), and used the same 3’ Cg CH1 reverse primer in each PCR reaction consisting of 3µl of cDNA, 1X Amplitaq Gold 360 Master Mix (Applied Biosystems), 125 nM of the reverse primer and 125 nM of the primer pool. The three IgH gamma pool amplicons were pooled and 2^nd^ round PCR reactions used the pooled primary PCR IgH amplicons as input. The second round PCR reactions mixes consisted of 1x AmpliTaq Gold 360 Master Mix, 500 nM 3’ IgGint Reverse Primer and 500 nM of respective nested Heavy Chain Forward Primer pool. Each round of IGH PCR was performed for 50 cycles at 94°C for 30 sec, annealing temp specified in NGS primer table for 30 sec, 72°C for 55 sec (1^st^ PCR) or 45 sec (2nd PCR) and a final 7 min extension cycle.

Light chain specific (Igλ or Igk) primer pools were used to amplify light chain specific immunoglobulin transcripts using previously published primers by two rounds of nested PCR for each light chain (see Supplemental for list of primers).

The primary PCR used 5 ul of cDNA as input and consisted of 500 nM of either CK-ext Reverse Primer (kappa) or CL-new-ext Reverse Primer (lambda) and either 2.5 µM Kappa Forward Primer Mix or 5 µM Lambda Forward Primer Mix and 1x AmpliTaq Gold 360 Master Mix. Nested PCR used 3 µL of the respective primary light chain amplicon with1x AmpliTaq Gold 360 Master Mix, 500 nM of either CK-int Reverse Primer (kappa) or CL-int Reverse Primer (lambda) and 500 nM of respective Nested Kappa/Lambda Chain Forward Primer. Each round of light chain PCR used the following amplification parameters 95°C for 5 min, 50 cycles of 95°C for 30 sec, Annealing temperature specified in primer table, for 30 sec and 72°C for 60 sec, followed by a final extension of 72°C for 7 min.

Nested PCR products for all chains were cleaned and normalized using the AxyPrep MAG PCR Normalizer Kit (Axygen). Concentrations post-normalization were confirmed with Qubit dsDNA High Sensitivity Assay Kit (Invitrogen) and diluted where necessary. Sample pools were constructed by taking 5 μL of each normalized PCR product from matching samples and combining together into one 1.5 mL tube. Each pool was mixed by pipetting, and then 5 μL of each pool were added into a 96-well plate, with each sample pool added to a different well. Sequencing libraries were constructed using the Nextera XT DNA Library Preparation Kit (Illumina) according to the manufacturer’s protocol. Indexed libraries were normalized, pooled and sequenced using (300 paired end, version 3 600 cycle kit, Illumina) on a MiSeq Illumina sequencer.

### Processing and BCR repertoire analysis

Paired-end sequencing reads were assembled by invoking IMMCANTATION/pRESTO AssemblePairs.py align. Details of BCR assembly have been published previously (Vander Heiden et al., 2014). Post-assembly filtering of the assembled reads was accomplished using IMMCANTATION/pRESTO FilterSeq.py, first by removing assembled reads with length less than 250 nt, and then by filtering out assembled reads with quality scores less than 30. The remaining assembled reads were written to FASTA files and submitted to IMGT/HighV-QUEST for analysis, including further filtering-out of reads determined to be unproductive (Lefranc et al., 2009).

### Single-cell BCR sequencing

Samples were single-cell sorted into 96-well PCR plates containing 10 µL of DNA Suspension Buffer (Teknova) with 1 mg/mL BSA (Sigma-Aldrich) per well. Plates were sealed and briefly centrifuged before being placed on dry ice. Plates were stored at −80°C at least overnight before proceeding. Plates were thawed on ice and spun at 800 x g for 1 minute to collect all liquid. Random primed cDNA synthesis was performed on each well using the SuperScript III Reverse Transcriptase kit with the following changes to the reaction mix: 5 mM DTT, 200 U SuperScript III and and 0.8 mM each dNTP mix. Amplification proceeded with 2 rounds of nested PCR targeting either the heavy chain, kappa chain and lambda chain (Supplemental) (Liao et al., 2009; Murugan et al., 2015). In each reaction, 2 μl of the cDNA product were used with 1x AmpliTaq Gold 360 Master Mix, 300 nM of chain-matched forward primer pool and 200 nM of chain-matched reverse primer. For heavy chain only, the reverse primer was in the form of two pooled primers that target either IgG or IgM sequences. Amplification parameters were 95°C for 5 min, 50 cycles of 95°C for 30 sec, 57°C (heavy and kappa) or 60°C (lambda) for 30 sec and 72°C for 55 sec, followed by a final extension of 72°C for 10 min.

Nested PCRs were run for each chain using 4 μl of unpurified first round PCR product. Three master mixes consisting of 1x AmpliTaq Gold 360 Master Mix and 200 nM chain-matched reverse primer were created and aliquoted to 96-well PCR plates. The first round PCR product was added to its chain-matched plate containing the nested PCR mix. Forward primers for each chain contained 12 nucleotide well IDs (heavy and kappa), the lambda chain forward primers contained the 8 nucleotide Chain specific plate IDs and reverse primers contained 8 nucleotide plate IDs (heavy and kappa) at their respective 5’ ends, and the lambda chain reverse primer contained 12 nucleotide well IDs. These barcoded forward primers were then spiked into the nested PCR plate of the matching chain at 200 nM. Amplification parameters were 95°C for 5 min, 50 cycles of 95°C for 30 sec, 57°C (heavy and kappa) or 60°C (lambda) for 30 sec and 72°C for 50 sec, followed by a final extension of 72°C for 10 min.

The wells from each corresponding heavy, kappa and lambda plate were the pooled by plate and bead cleaned using 0.6X HighPrep™ PCR Clean-up System SPRI magnetic beads (MagBio Genomics). Cleaned amplicons were indexed using the Qiaseq 1 step Amplicon DNA library kit (Qiagen) per manufacturer’s instructions. Each pooled and indexed sample was quantified and normalized to 4 nM using Kapa Biosystems Illumina library quantification kit per the manufacturer’s instructions. Indexed and normalized samples were then pooled and sequenced using (300 paired end, version 3, 600 cycle kit, Illumina) on a MiSeq Illumina sequencer.

### Processing and single-cell BCR sequencing analysis

Single-cell read sequences have a number of attributes, including the originating plate and well IDs, as well as information about the particular primer associated with a given reaction within a well (for heavy chains this is the “VREG” identifier while for light chains it is the “CPRIMER” attribute). The (plate, well, primer) triples serve to partition the read sequences into groups sharing the same attributes, and each of these read groups is then independently subjected to the following filtering steps:

(1) Retain a read sequence only if its duplicate count is ≥ 2,

(2) Sort the reads within a given group into descending order of read-sequence duplicate count. Note the V gene associated with the most frequently-occurring read sequence in the group and call its duplicate-count COUNT_MAX and its v-gene VGENE_MAX (the v-gene, including allele, is computed via IMGT/HighV-QUEST).

(3) Consider the other read sequences in the sorted list whose duplicate counts are greater than or equal to COUNT_MAX / 2. If any of these sequences’ V genes differ from VGENE_MAX then remove the entire group of read-sequences from further consideration. Otherwise, retain the read-sequences that have duplicate counts greater than or equal to COUNT_MAX / 2 and have common V gene VGENE_MAX.

Step 1 of filtering resulted in 171,010 read sequences with duplicate counts ≥ 2 and Steps 2 and 3 further reduced this set to 11,758 read sequences.

In an effort to eliminate the effects of possible cross contamination, after the above filtering steps had been applied to all read sequence groups, we constructed a mapping of the filtered set’s read sequences to the subjects that generated them. If a given read sequence was associated with more than one subject then all read sequence groups sharing that read sequence were filtered out (applying this shared-subject filtering step resulted in 6099 read sequences), and we proceeded to the next step of constructing a single consensus sequence for each group from these filtered retained sequences. Read sequence clustering and consensus resulted in 2449 sequences associated with 1911 wells, 339 of which had light chains paired with heavy chains.

### Statistical analyses

Paired comparisons (e.g., frequencies of V genes between the gp120^+^ and gp120^-^ repertoires) were carried out using the Wilcoxon signed-rank (WSR) test. Associations between paired variables were measured using Spearman’s rank correlation coefficients. P-values adjustment for multiple comparisons was done using Benjamini and Yekutieli’s procedure (Benjamini and Yekutieli, 2001). Adjusted p-values smaller than 0.10 were considered statistically significant. The diversity of V genes within individuals that encoded the heavy, kappa, and lambda chains of gp120^+^ and gp120^-^ B cells were compared using three diversity indices: the richness index, defined as the number of distinct V gene families observed in gp120^+^ and gp120^-^ samples; Shannon diversity, which is based on the geometric mean of the proportional abundances of observed V gene frequencies; and Simpson diversity index, which estimates the probability that two read sequences sampled at random without replacement belong to the same V gene family. Unlike the richness and Shannon indices, a *lower* Simpson diversity indicates a *higher* diversity. These indices were compared between classes (gp120^+^ and gp120^-^ B cells) using linear mixed models (LMM) adjusting for log-read count and class, and including subject-specific random intercepts. We used Jaccard’s beta diversity index to evaluate the overlap between the pool of V genes used by the repertoire of different vaccine-recipients to encode their gp120^+^ and gp120^-^ heavy and light chains. The Jaccard index between two repertoires was calculated as *J*_12_ = *C*_12_/(*C*_12_ + *U*_1_ + *U*_2_) where *C*_12_ denotes the number of genes shared by the two repertoires and *U*_1_ and *U*_2_ are the numbers of V genes unique to the first and second repertoire, respectively.

To assess evolution of BCR due to somatic hypermutation and antigen-driven selection during germinal center reactions, we calculated the Hamming amino acid distance between the V region of read sequences and their germline V gene. To examine convergence of the vaccine-induced repertoire toward *V_H_*1-2*02-restricted bnAbs, we computed distances between read sequences and 5 such HIV-1 bnAbs: VRC01, IOMA, N6, DH270.1, and VRC-PG04. Distances were separately computed by segment of the V region: FR1-3, and CDR1-2. Calculations of Hamming distances were restricted to read sequences with complete coverage of the FR1-FR3 regions.

To identify CDRH3 clonotypes, we first computed pairwise distances between AA CDRH3 read sequences using the Levenshtein distance. We next performed single-linkage clustering to identify BCR with similar CDRH3 a.a. sequences. The threshold used to identify the minimum distance between clusters was determined by inspecting the distribution of the pairwise distances. A sensitivity analysis was carried out to evaluate the impact of the value of the threshold on the results.

## Supporting information

Supplemental Primer Tables

Supplemental vgene Frequencies

## Data Availability

All data is included in the manuscript or is accessible from the corresponding authors.

## Acknowledgements

We like to thank the HVTN 100 protocol chairs, protocol teams and participants for making this study possible.

