## Supplemental Figures for "Shared HIV envelope-specific B cell clonotypes induced by a pox-protein vaccine regimen"

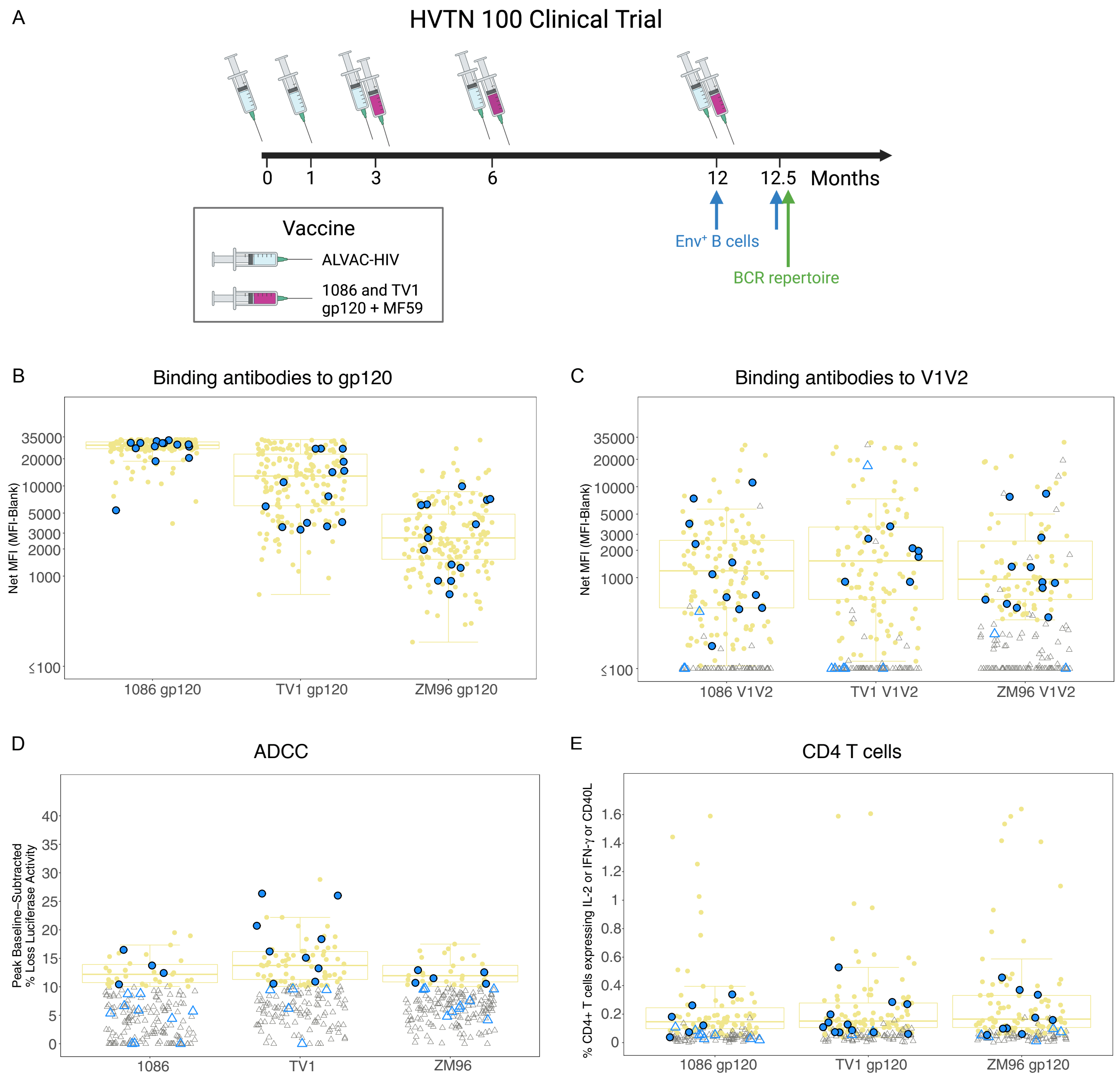


**Figure S1. HVTN 100 clinical trial and immunogenicity data at month 12.5.** A) Trial schema for the HVTN 100 clinical trial indicating the vaccination schedule and Env-specific B cell phenotyping and BCR repertoire analysis timepoints. Immune responses at month 12.5 to vaccine-matched antigens for the B cell analysis subset (n=14) in blue overlayed on the entire vaccinated cohort in yellow (n=185): Binding antibody responses to vaccine-matched B) gp120 and C) V1V2 proteins. D) Antibody-dependent cellular cytotoxicity (ADCC) activity of serum as measured by luciferase-based cytotoxicity assay. E) CD4 T cell responses (IL-2, IFN-γ, or CD40L) to vaccine-matched gp120 peptide pools. Colored circles indicate statistically “positive” response whereas open triangles indicate non-response.

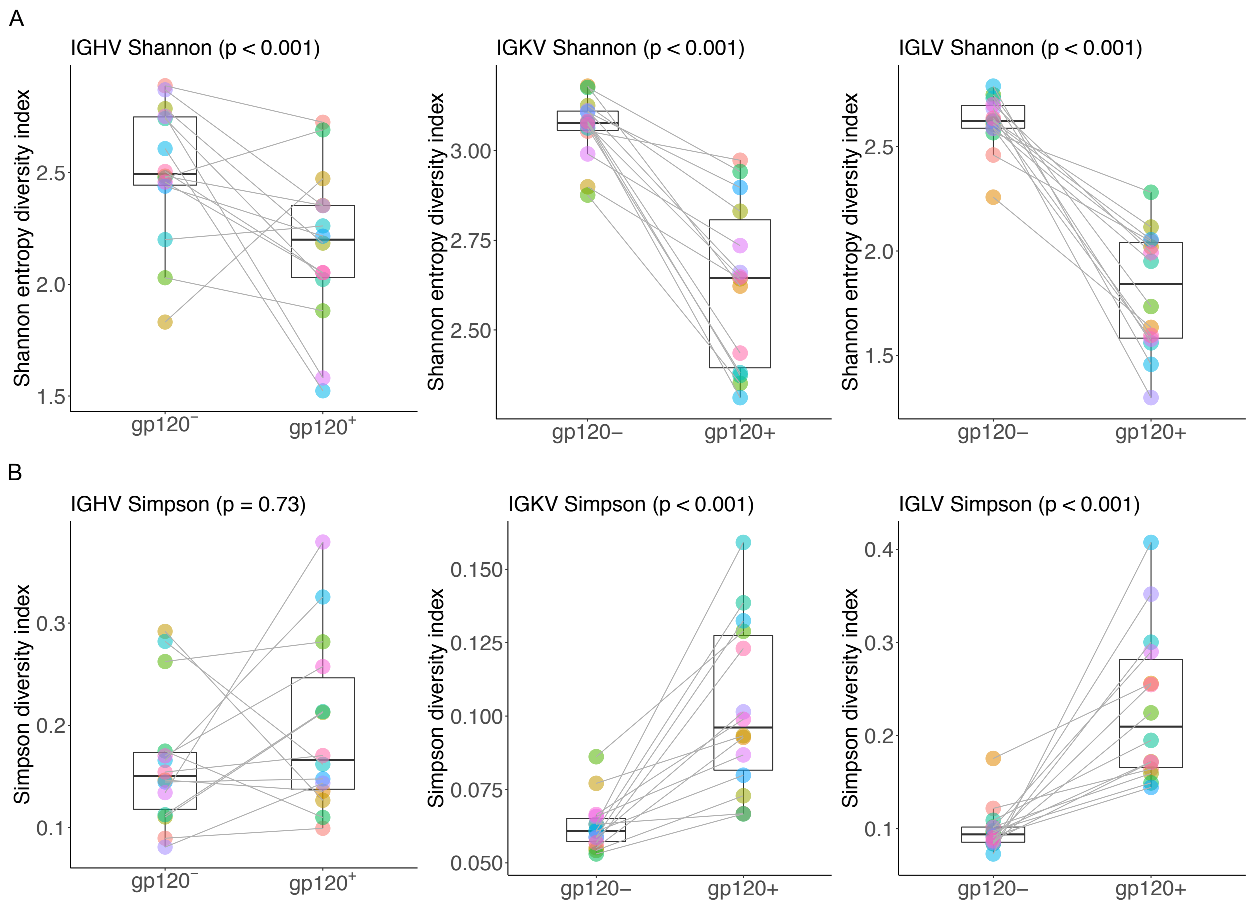


**Figure S2. Diversity of $V_H$, $V_K$ and $V_L$ s for the gp120$^-$ and gp120$^+$ repertoires as calculated by the A) Shannon or B) Simpson indices.**

A

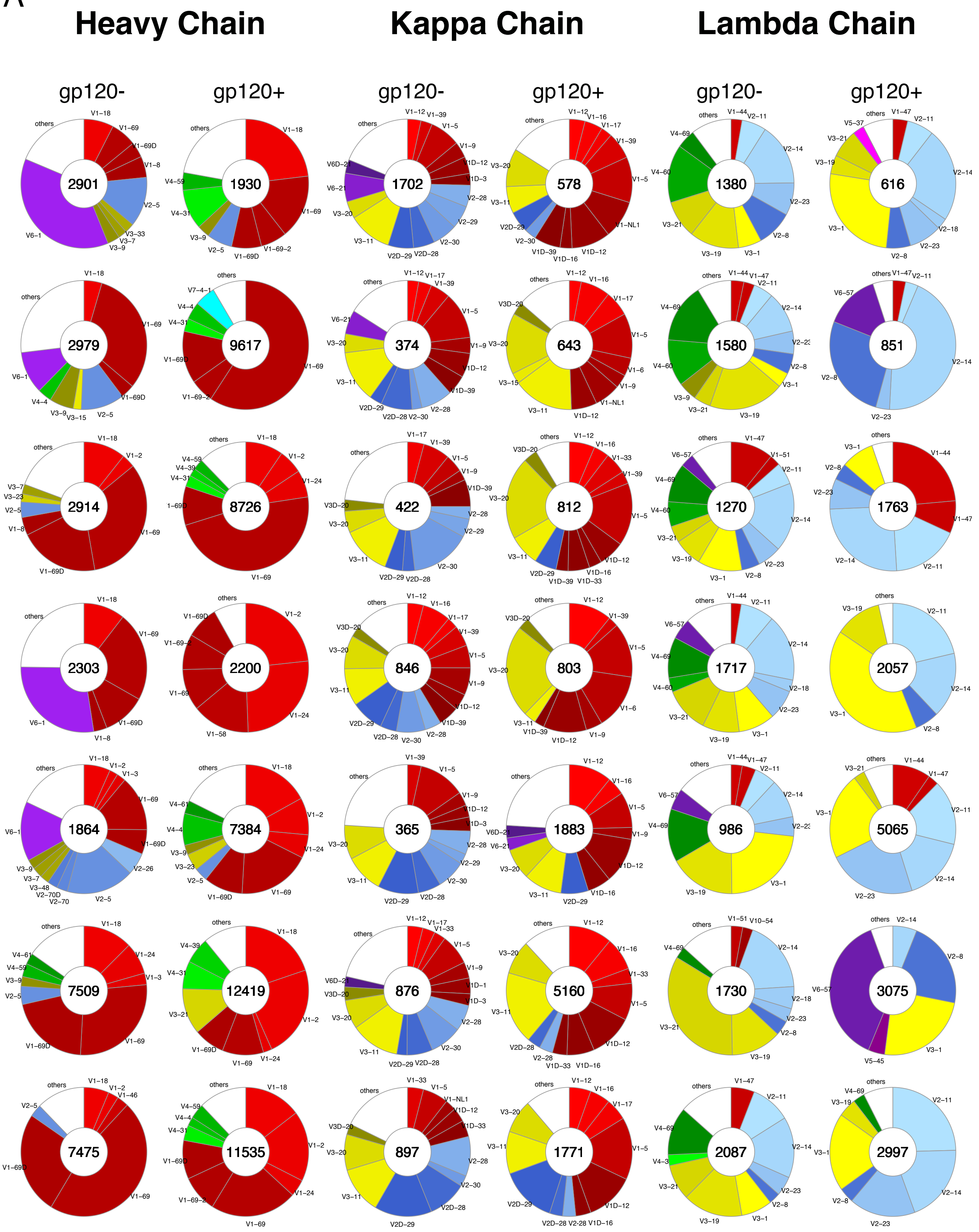
Heavy Chain
Kappa Chain
Lambda Chain
gp120-
gp120+
gp120-
gp120+
gp120-
gp120+
2901
1930
1702
578
1380
616
2979
9617
374
643
1580
851
2914
8726
422
812
1270
1763
2303
2200
846
803
1717
2057
1864
7384
365
1883
986
5065
7509
12419
876
5160
1730
3075
7475
11535
897
1771
2087
2997

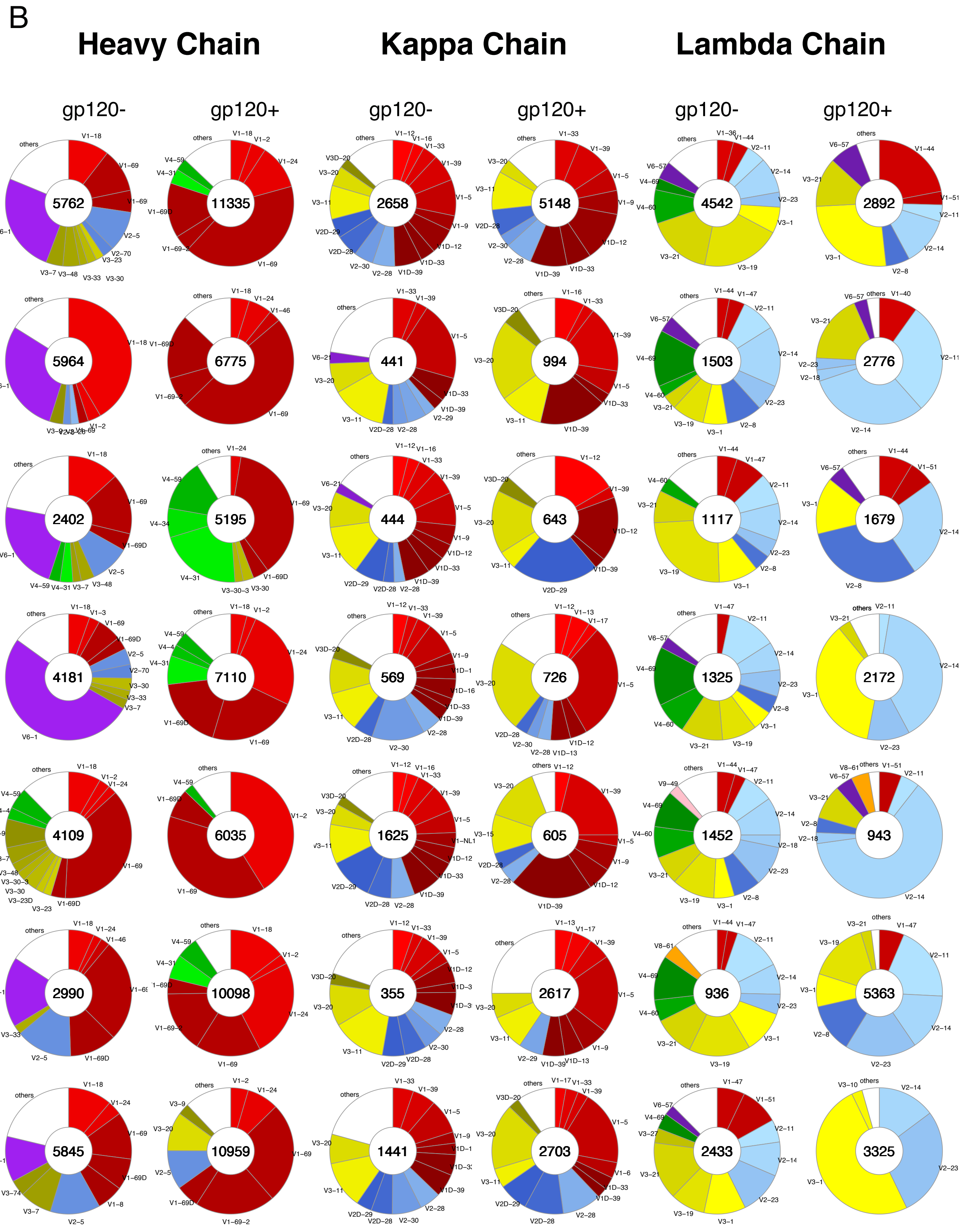
B
Heavy Chain
Kappa Chain
Lambda Chain
gp120-
gp120+
gp120-
gp120+
gp120-
gp120+
5762
11335
2658
5148
4542
2892
5964
6775
441
994
1503
2776
2402
5195
444
643
1117
1679
4181
7110
569
726
1325
2172
4109
6035
1625
605
1452
943
2990
10098
355
2617
936
5363
5845
10959
1441
2703
2433
3325

**Figure S3. Proportions of $V_H$, $V_K$ and $V_L$ sequences encoded by specific V genes for the gp120- and gp120+ repertoires, by subject (one subject per row): A) subjects 1-7 and B) 8-14.** The total number of sequences per repertoire is indicated in the center of the pie chart. Each V gene family (e.g., VH1, VH2...) is highlighted by a specific color that is identical across chains (e.g., all genes in the VH1, VL1 and VK1 families are shown in red); different V gene segments within a V gene family (e.g., VH1-2) are represented by different shades of the same color.

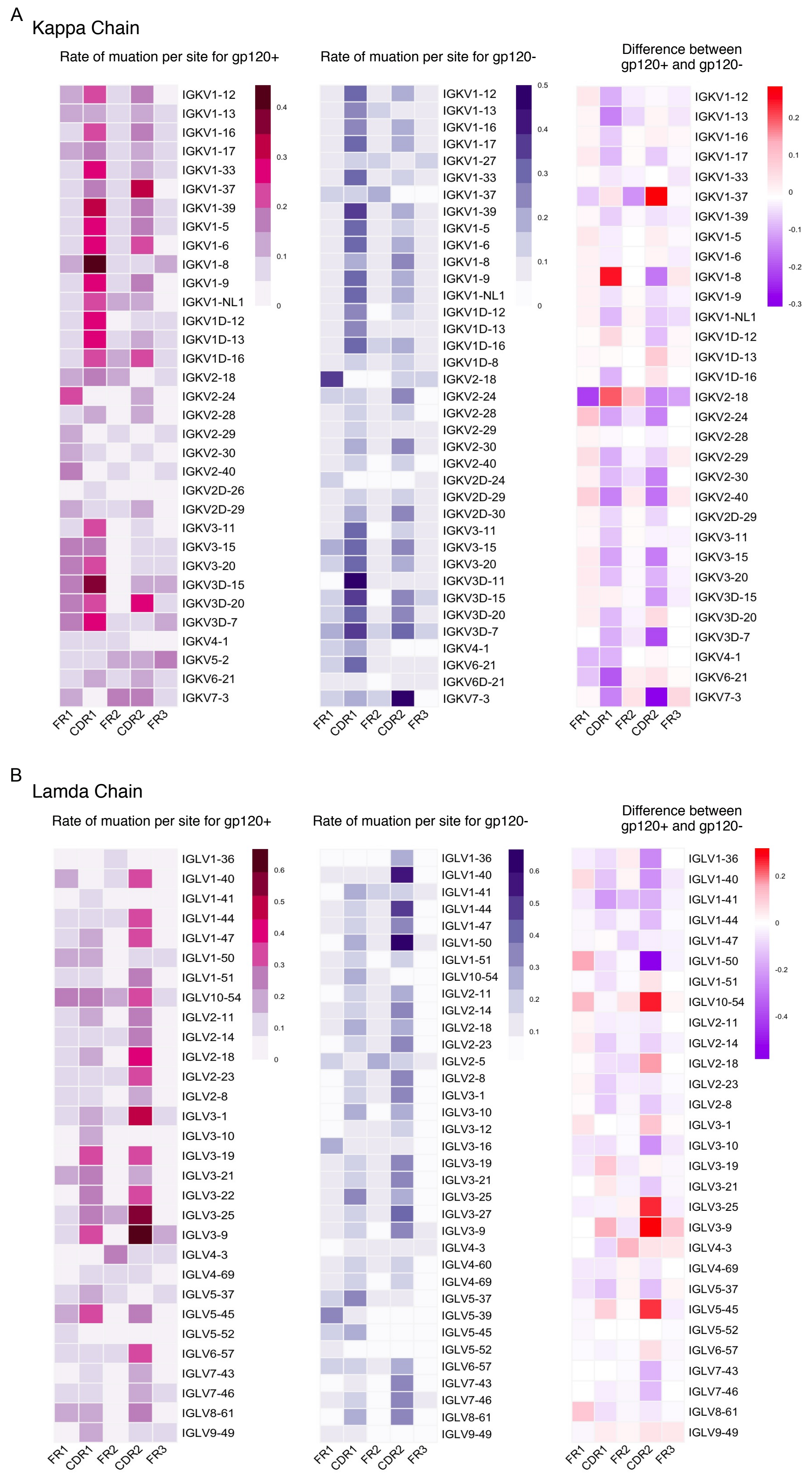

A
Kappa Chain
Rate of muation per site for gp120+
Rate of muation per site for gp120-
Difference between gp120+ and gp120-
FR1
CDR1
FR2
CDR2
FR3
IGKV1-12
IGKV1-13
IGKV1-16
IGKV1-17
IGKV1-27
IGKV1-33
IGKV1-37
IGKV1-39
IGKV1-5
IGKV1-6
IGKV1-8
IGKV1-9
IGKV1-NL1
IGKV1D-12
IGKV1D-13
IGKV1D-16
IGKV1D-8
IGKV2-18
IGKV2-24
IGKV2-28
IGKV2-29
IGKV2-30
IGKV2-40
IGKV2D-24
IGKV2D-26
IGKV2D-29
IGKV2D-30
IGKV3-11
IGKV3-15
IGKV3-20
IGKV3D-11
IGKV3D-15
IGKV3D-20
IGKV3D-7
IGKV4-1
IGKV5-2
IGKV6-21
IGKV6D-21
IGKV7-3
0.5
0.4
0.3
0.2
0.1
0
-0.1
-0.2
-0.3
B
Lamda Chain
Rate of muation per site for gp120+
Rate of muation per site for gp120-
Difference between gp120+ and gp120-
FR1
CDR1
FR2
CDR2
FR3
IGLV1-36
IGLV1-40
IGLV1-41
IGLV1-44
IGLV1-47
IGLV1-50
IGLV1-51
IGLV10-54
IGLV2-11
IGLV2-14
IGLV2-18
IGLV2-23
IGLV2-5
IGLV2-8
IGLV3-1
IGLV3-10
IGLV3-12
IGLV3-16
IGLV3-19
IGLV3-21
IGLV3-22
IGLV3-25
IGLV3-27
IGLV3-9
IGLV4-3
IGLV4-60
IGLV4-69
IGLV5-37
IGLV5-39
IGLV5-45
IGLV5-52
IGLV6-57
IGLV7-43
IGLV7-46
IGLV8-61
IGLV9-49
0.6
0.5
0.4
0.3
0.2
0.1
0
-0.2
-0.4

**Figure S4. Rate of mutation among gp120+ and gp120- kappa and lambda sequences.** Mean amino acid (a.a.) mutation rates per site in the respective framework (FR) or complementary determining (CDR) regions of the A) $V_K$ and B) $V_L$ sequences split out by V gene for the gp120+ or gp120- repertoires, followed by the difference in a.a. mutation rate between the gp120+ and gp120- sequences per $V_K$ or $V_L$ region. In the heatmap that shows the difference between mutation rates in the gp120+ and gp120- repertoires, color of heatmap indicates higher (red) or lower (purple) mutation rates in the gp120+ than in the gp120- repertoire.
